# Multimodal associations between posterior hippocampus glutamate metabolism, visual cortex connectivity, and intrusive trauma reexperiencing symptoms

**DOI:** 10.1101/2025.01.27.25320595

**Authors:** Kevin J. Clancy, Xi Chen, Xiaopeng Song, Tao Song, Shuqin Zhou, Eylül Akman, Caroline Ostrand, Boyu Ren, Fei Du, Isabelle M. Rosso

## Abstract

**Objective:** Hippocampal dysfunction is implicated in posttraumatic stress disorder (PTSD), particularly intrusive reexperiencing symptoms, and may be mediated by glutamatergic excitotoxicity. Markers of glutamate dysfunction (higher glutamate to N-acetyl aspartate levels; Glu/NAA) in the hippocampus (HPC) have been linked to reexperiencing symptoms. However, the HPC demonstrates heterogeneity along its anterior-posterior axis, with different functional connectivity patterns and PTSD symptom associations, motivating investigations into glutamate metabolism in anterior and posterior HPC subregions (a/pHPC).

**Methods:** 121 symptomatic trauma-exposed adults (93 female) with current trauma reexperiencing symptoms completed magnetic resonance spectroscopy and resting-state functional magnetic resonance imaging to examine the regional specificity of HPC Glu/NAA associations with reexperiencing, and the link to a/pHPC functional connectivity. PTSD symptoms were assessed with the Clinician-Administered PTSD Scale for DSM-5.

**Results:** Reexperiencing symptom severity was associated with greater Glu/NAA in the pHPC, but not aHPC. pHPC Glu/NAA was further linked to stronger functional connectivity between the pHPC and visual cortex (VC), which in turn correlated with more severe reexperiencing symptoms. This strengthened pHPC-VC connectivity explained the shared variance between pHPC Glu/NAA and reexperiencing severity, suggesting dysregulated glutamate metabolism in the pHPC may contribute to reexperiencing symptoms through functional connectivity with the VC.

**Conclusions:** These findings replicate prior work linking HPC Glu/NAA to trauma reexperiencing symptoms and provide novel evidence this association may be specific to the pHPC and mediated by its functional connectivity with the VC. This multimodal investigation supports translational models of glutamatergic dysfunction in trauma-related disorders and highlights new targets for pharmacological and neuromodulatory interventions.

## INTRODUCTION

The intrusive reexperiencing of a traumatic event as memories, nightmares, flashbacks, or reactivity to trauma reminders, is common among individuals exposed to trauma and represents a central set of symptoms in posttraumatic stress disorder (PTSD) [1]. Beyond PTSD, these reexperiencing symptoms predict the onset, maintenance, and severity of different transdiagnostic psychiatric difficulties following trauma exposure [2,3]. Additionally, these symptoms independently cause clinically significant distress and impairment in subthreshold presentations [4]. As such, they are positioned as central therapeutic targets to address the growing impacts of trauma-related psychiatric distress and impairment [4].

Intrusive reexperiencing symptoms are memory-related phenomena, with proposed neurobiological substrates predominantly focused on the hippocampus [5]. The hippocampus is key in forming and retrieving episodic memories [6] and distinguishing between safe and dangerous contexts [7]. Neurobiological investigations of trauma-related disorders consistently demonstrate hippocampal atrophy and dysfunction, linking these changes to fear learning and memory models of PTSD [8,9]. Smaller volumes and hypoactivity of the hippocampus may hinder the ability to learn and remember safety cues in people with PTSD [5]. These deficits may contribute to exaggerated activations of learned fear memories and difficulties regulating amygdala-mediated arousal to threat cues.

Mechanistic accounts of hippocampal dysfunction in trauma-related disorders implicate glutamatergic excitotoxicity, whereby elevated glutamate (Glu), the predominant excitatory neurotransmitter, result in neuronal dysfunction and injury [10]. Extreme stress increases glutamatergic signaling, which may mediate stress-induced atrophy of hippocampal neurons [10]. This glutamatergic neurotoxicity is particularly pronounced following repeated or prolonged stress [11], and has been linked to deficits in learning and memory and alterations of hippocampal networks [12].

*In vivo* investigations of glutamatergic neurotoxicity have positioned the ratio of Glu to N-acetyl aspartate (NAA), a marker of neuronal integrity and metabolism, as a proxy measure of excitotoxicity [13,14]. Magnetic resonance spectroscopy (MRS) allows researchers to reliably estimate regional brain Glu and NAA levels *in vivo* in humans. Lower hippocampal NAA levels are consistently observed in PTSD, aligning with findings of hippocampal dysfunction and atrophy [15,16]. MRS studies of neurological disorders associated with excitotoxicity, such as epilepsy and amyotrophic lateral sclerosis, have demonstrated higher Glu/NAA ratios, reflecting elevated excitatory metabolism (higher Glu) and compromised neuronal integrity or function (lower NAA) [14,17]. In the first MRS examination of hippocampal Glu metabolism in PTSD, our group demonstrated elevated Glu and Glu/NAA ratios in the right hippocampus of PTSD patients compared to asymptomatic trauma-exposed controls, which scaled with reexperiencing symptom severity [13]. This provided foundational human evidence for the glutamate excitotoxic model of hippocampal dysfunction in PTSD, particularly in relation to memory-based symptoms of intrusive reexperiencing.

While prior PTSD studies have examined the hippocampus as a unitary structure, accumulating evidence highlights its functional heterogeneity along the anterior-posterior (a-p) axis [18,19]. The posterior hippocampus (pHPC) supports the retrieval of visuospatial memory details for reconstructing vivid scenes of past events [6,20,21]. Conversely, the anterior hippocampus (aHPC) retrieves the affective or schematic “gist” of memories, integrating sensory-perceptual and cognitive-affective features into an overarching memory construct [22]. This specialization is reflected in distinct functional connectivity patterns [21]: the aHPC is primarily coupled with the default mode network, frontal cortical structures, and the amygdala – areas involved in cognitive and affective processes. In contrast, the pHPC is more strongly coupled with the visual cortex and multimodal sensory regions that support sensory-perceptual processes, like mental imagery.

Investigations into the phenomenology of reexperiencing symptoms highlight their sensory-perceptual vividness that contributes to a strong sense of reliving the trauma in the “here-and-now” [23]. Given the pHPC’s role in supporting sensory-rich details of memory, these phenomenological accounts suggest its critical involvement in reexperiencing symptoms, perhaps independently of the aHPC [24]. Recent findings from our group have linked strengthened dynamic coupling between the pHPC and visual cortex to the reliving qualities of intrusive trauma memories [25]. This suggests that pHPC interactions with visual systems as a candidate mechanism for reexperiencing symptoms.

This study aimed to replicate our prior findings linking elevated Glu/NAA in the right hippocampus to intrusive reexperiencing symptoms in PTSD patients [13], and to extend them by investigating their regional specificity within the right aHPC and pHPC. Consistent with our hypothesis that these effects would be specific to the pHPC, we further investigated how pHPC Glu/NAA related to functional connectivity patterns demonstrate a potential candidate mechanism through which pHPC excitotoxicity may contribute to reexperiencing symptoms. Specifically, we hypothesized that elevated Glu/NAA would be associated with stronger pHPC–visual cortex connectivity. Finally, we examined whether trauma load and PTSD diagnosis moderated these associations, given the well-established effects of chronic stress and evidence this may emerge only in those with a PTSD diagnosis.

## METHODS

### Participants and Procedures

We enrolled 121 trauma-exposed adults from the community, aged 18-65 years, who were included if they reported experiencing at least two trauma-related intrusive memories per week, as part of a larger study [25,26]. Study procedures were approved by the Mass General Brigham Human Research Committee, and all participants provided written informed consent. Detailed selection criteria are provided in the Supplement.

Participants completed the Clinician-Administered PTSD Scale for DSM-5 (CAPS-5) and magnetic resonance imaging (MRI), which included resting-state functional MRI (rs-fMRI) to measure functional connectivity and magnetic resonance spectroscopy (MRS) to assess Glu/NAA ratios in the aHPC and pHPC. MRS data from the aHPC were excluded for 7 participants due to poor quality (see Supplement). rs-fMRI data were not acquired from 2 participants due to technical difficulties and were excluded from 12 participants due to confounds or poor quality (falling asleep in 2 participants, excessive motion in 10 participants).

### Interview and self-report measures

#### Clinician-Administered PTSD Scale for DSM-5 (CAPS-5)

The CAPS-5 [27] was administered by doctoral-level clinicians to confirm that all participants endorsed a Criterion A trauma and assess severity of all PTSD symptoms (Clusters B-D). The total score was calculated as the sum of all 20 symptom scores (each rated on five-point Likert scale from 0 = Absent to 4 = Extreme), and the intrusive reexperiencing symptom cluster score as the sum of scores for the five symptoms in that cluster (B1-B5). Table 1 shows that most participants met diagnostic criteria for PTSD in this sample.

**Table 1.** Demographic and clinical characteristics. Mean ± standard deviation or N (%).

| Total Sample (n = 121) |  |
| --- | --- |
| Age (years) | 33.7 ± 11.0 |
| Sex assigned at birth |  |
| Female | 93 (77%) |
| Male | 28 (23%) |
| Gender |  |
| Woman | 79 (65%) |
| Man | 29 (24%) |
| Gender fluid/non-binary | 13 (11%) |
| Race |  |
| White | 87 (72%) |
| Multiracial | 14 (12%) |
| Black | 7 (6%) |
| Asian | 8 (7%) |
| Unknown or Not Reported | 5 (4%) |
| Ethnicity |  |
| Non-Hispanic | 113 (93%) |
| Hispanic | 8 (7%) |
| Current Psychotropic Medication | 56 (46%) |
| Antipsychotic | 8 (7%) |
| Antidepressant | 45 (37%) |
| Mood Stabilizer | 16 (13%) |
| Sedative/Hypnotic | 20 (17%) |
| Stimulant | 19 (16%) |
| CAPS-5 Cluster B Severity<br>(Reexperiencing Symptoms) | 9.7 ± 3.3 |
| CAPS-5 Total Score | 34.6 ± 11.0 |
| CAPS-5 PTSD Diagnosis | 98 (81%) |
| TLEQ Trauma Load | 29.8 ± 17.1 |

Sex assigned at birth, gender identity, race, and ethnicity data were acquired via participant self-report.
CAPS-5 = Clinician-Administered PTSD Scale for DSM-5; TLEQ = Traumatic Life Events Questionnaire.

#### Traumatic Life Events Questionnaire (TLEQ)

The TLEQ is a self-report assessment of lifetime traumas [28], each rated for occurrence (yes/no) and frequency. We derived a “trauma load” score by summing the total number of traumatic events experienced over participants’ lifetimes.

### Functional magnetic resonance imaging

Eyes-open resting-state fMRI data (13 minutes, 976 volumes) were acquired on a 3T Siemens Prisma scanner with a 64-channel head coil using the Human Connectome Project (HCP) Lifespan protocol. In addition, T1-weighted 3D MPRAGE anatomical images were obtained using the HCP 0.8mm resolution sequence. Participants were excluded if their mean framewise displacement (FD) exceeded 0.5 mm or greater than 20% of volumes exceeded FD = 0.5mm (n = 10). Scan sequences and preprocessing steps are detailed in the Supplement.

Cleaned timeseries were extracted from the aHPC and pHPC regions of interest (ROIs), defined by a study-specific probabilistic template [26]. Whole-brain seed-to-voxel correlations were performed using the resulting a/pHPC seeds to compute whole-brain functional connectivity maps.

### Magnetic resonance spectroscopy

Single-voxel proton MRS was acquired from two voxels (each 20mm x 10mm x 20mm) positioned in the right aHPC and pHPC (**Figure 1A**). Given previous findings associating reexperiencing symptoms with Glu/NAA in the right, but not left, hippocampus [13], data acquisition was constrained to the right hippocampus. A semi-LASER sequence [29] with water saturation by VAPOR approach (64 averages) was used to optimally measure Glu and NAA concentrations. Unsuppressed water signals from the same voxels and using the same sequence were collected for frequency, phase, and eddy-current correction and served as the external reference for metabolite quantification, controlling for proportions of gray matter, white matter, and cerebrospinal fluid proportions, as well as relaxation effects i.e. partial volume corrections [30,31]. Details and representative spectra are presented in the Supplement.

**Figure 1.**
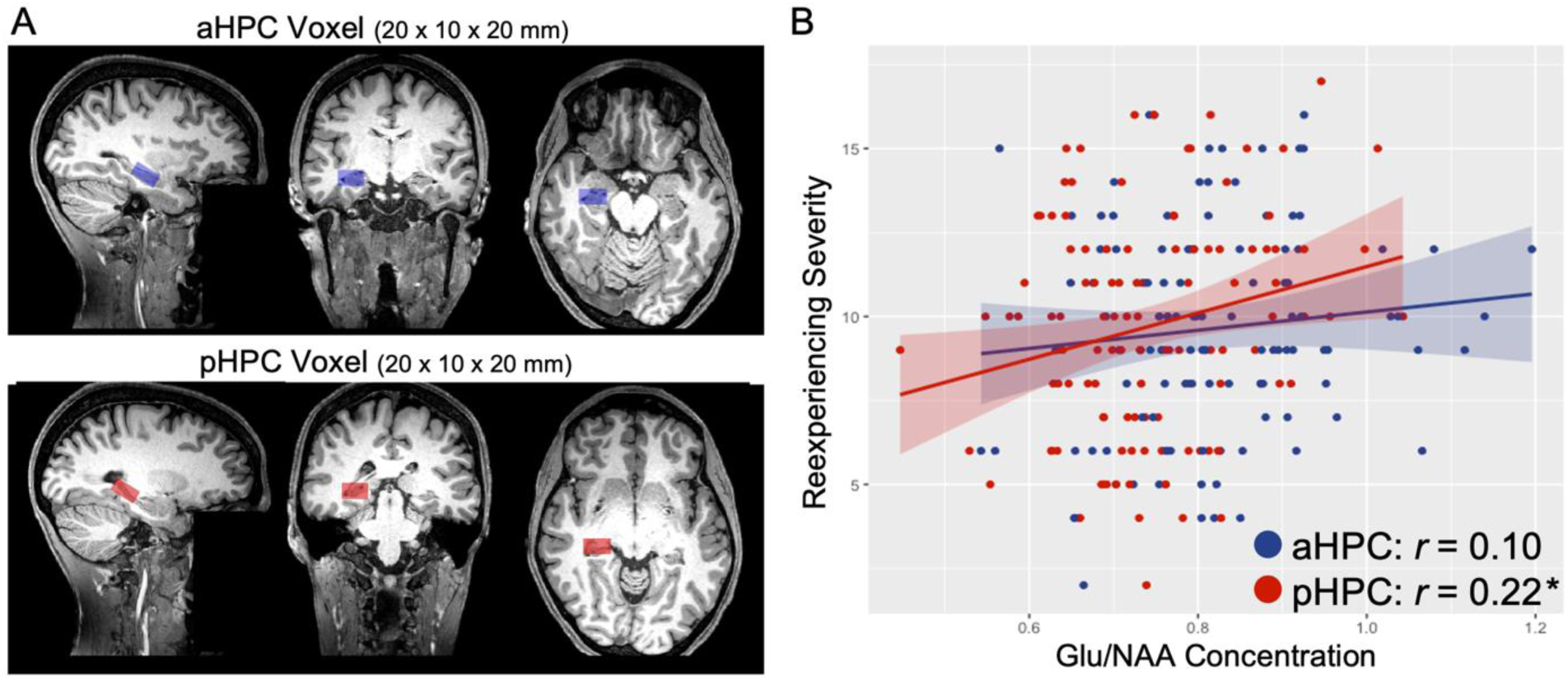
Relationships of anterior hippocampus (aHPC) and posterior hippocampus (pHPC) Glu/NAA with intrusive reexperiencing symptoms. A) Representative MRS voxels in the aHPC (blue) and pHPC (red). B) Association between reexperiencing symptom severity and Glu/NAA in the pHPC but not aHPC. \**p* < 0.05. Note: Glu = glutamate; MRS = magnetic resonance spectroscopy; NAA = N-acetyl-aspartate.

### Statistical Analyses

Pearson correlations (two-tailed) assessed relationships between CAPS-5 reexperiencing symptom severity and Glu/NAA ratios in the aHPC and pHPC. For significant correlations, follow-up linear regressions included age, sex assigned at birth, and current medication status (yes/no) as covariates to test the sensitivity of these associations to common covariates. Sex assigned at birth was significantly associated with reexperiencing symptoms (females > males; t = 2.26, p = 0.028).

To examine their associations with aHPC and pHPC functional connectivity, Glu/NAA from these regions was regressed onto whole-brain a/pHPC connectivity maps. Whole-brain results were analyzed using an uncorrected height threshold of p < 0.005 and a corrected cluster-size threshold of p < 0.05 FWE. To determine whether the connectivity patterns associated with Glu/NAA were associated with reexperiencing symptoms, subject-level connectivity estimates were extracted and correlated with reexperiencing severity scores. Whole-brain connectivity analyses were performed to demonstrate the robustness and spatial specificity of these effects by regressing reexperiencing severity onto a/pHPC-seeded connectivity maps.

In the presence of three-way correlations, a cross-sectional mediation model tested whether HPC connectivity explained the shared variance between HPC Glu/NAA and reexperiencing severity. Competing models were tested by swapping HPC Glu/NAA and connectivity as the predictor and mediator, recognizing that the cross-sectional study design precludes conclusions about directionality or causality.

Finally, the effect of total lifetime trauma exposure (TLEQ Trauma Load) and PTSD diagnosis on all variables of interest were examined through linear regressions (for TLEQ Trauma Load) and group contrasts (PTSD vs. no PTSD). Additionally, HPC Glu/NAA and connectivity models demonstrating significant effects were re-run with trauma load or PTSD diagnosis entered as a moderator of the effect. To account for the inherent relationship between age and lifetime trauma exposure, age was regressed out from TLEQ Trauma Load.

## RESULTS

### Regional specificity of Glu/NAA and intrusive reexperiencing symptoms

Correlating reexperiencing symptoms with pHPC and aHPC Glu/NAA, separately, revealed an association for the pHPC (r(119) = 0.22, p = 0.018, 95% CI = [0.04, 0.38]) but not aHPC (r(112) = 0.10, p = 0.292, 95% CI = [-0.09, 0.28]), such that higher pHPC Glu/NAA correlated with more severe reexperiencing symptoms (**Figure 1B**). This association remained significant after controlling for age, sex, and medication status (p = 0.013).

### pHPC Glu/NAA associations with functional connectivity

Regressing pHPC Glu/NAA onto whole-brain pHPC-seeded functional connectivity maps revealed a significant cluster in the visual cortex (cuneus; peak = -4, - 80, 24; t = 4.14; k = 322, cluster pFWE = 0.001), such that higher pHPC Glu/NAA was associated with stronger pHPC-visual cortex connectivity (**Figure 2A**). Notably, this cluster overlapped with a visual cortex cluster that demonstrated preferential connectivity with the pHPC versus aHPC (**Figure 2B**). No other clusters emerged. This effect persisted when controlling for age, sex, and medication (cluster pFWE = 0.001). No associations were seen with the aHPC.

**Figure 2.**
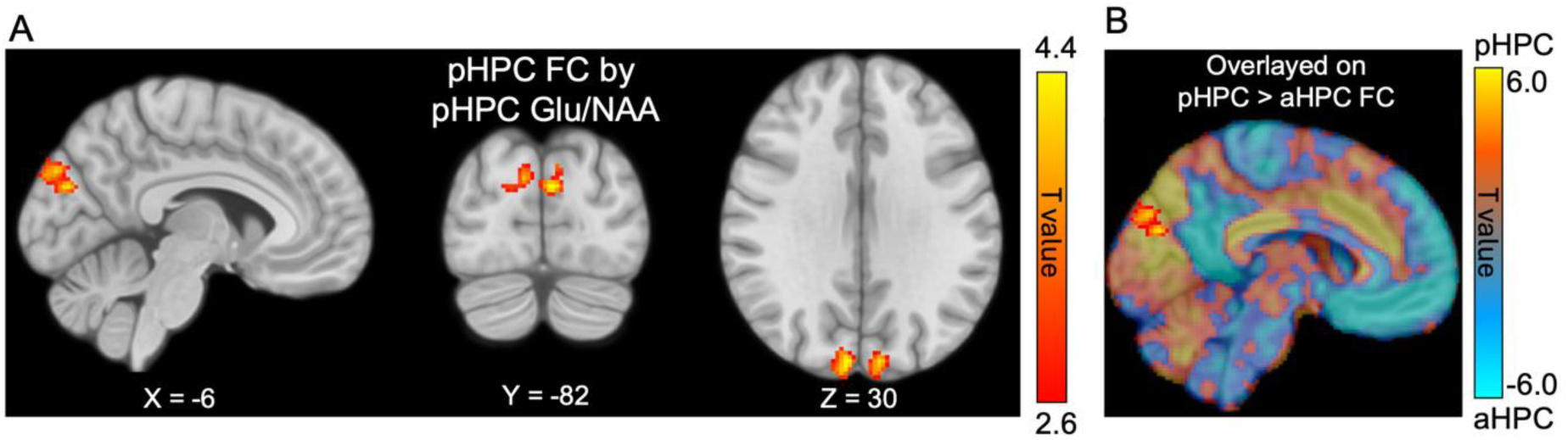
Association between posterior hippocampus (pHPC) Glu/NAA and functional connectivity (FC). A) pHPC Glu/NAA was associated with increased FC between the pHPC and a cluster in the visual cortex (cuneus; peak = -4, -80, 24). Display threshold set at voxel-wise p < 0.005, k = 20. B) pHPC Glu/NAA cluster overlayed on a paired t-test contrast map of pHPC versus aHPC whole-brain FC maps, demonstrating preferential pHPC connectivity with the visual cortex, which overlapped with the cluster associated with Glu/NAA ratios. Note: Glu = glutamate; NAA = N-acetyl-aspartate.

### pHPC functional connectivity associations with reexperiencing symptoms

Connectivity within the identified pHPC-visual cortex circuit was also associated with reexperiencing severity (r(105) = 0.30, p = 0.002, 95% CI = [0.12, 0.46]; **Figure 3A**). Regressing symptoms onto whole-brain pHPC-seeded functional connectivity maps revealed two significant clusters in the visual cortex: one in the cuneus overlapping with the region associated with pHPC Glu/NAA (peak = -6, -96, 18; t = 4.68; k = 317; cluster pFWE = 0.001) and another spanning the calcarine sulcus into the right lingual gyrus (peak = 8, -96, -4; t = 3.82; k = 287; cluster pFWE = 0.002; **Figure 3B**). No other clusters emerged.

**Figure 3.**
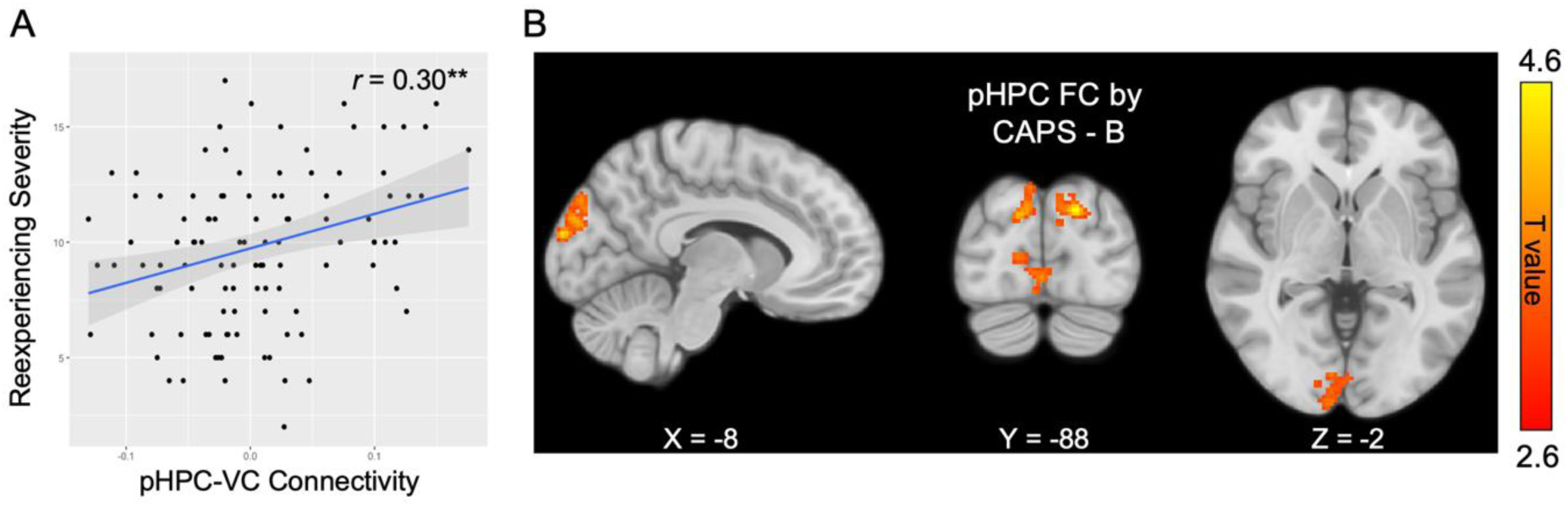
Association between posterior hippocampus (pHPC) functional connectivity and reexperiencing symptom severity. A) Significant correlation between reexperiencing symptom severity and functional connectivity within the pHPC-visual cortex circuit associated with pHPC Glu/NAA. B) Whole-brain regression of reexperiencing symptom severity (CAPS-B score) on pHPC-seed functional connectivity maps, demonstrating the spatial specificity of these effects to the visual cortex. Display threshold set at voxel-wise p < 0.005, k = 20. \*\**p* < 0.005. Note: CAPS-B = Clinician-Administered Posttraumatic Stress Disorder Scale, Cluster B score; Glu = glutamate; NAA = N-acetyl-aspartate.

### Mediation Analyses

Examining the variance explained across the associations between symptoms, Glu/NAA, and functional connectivity revealed that pHPC-visual cortex connectivity mediated the association between pHPC Glu/NAA and reexperiencing severity (indirect effect: B = 0.54, 95% CI = [0.06, 2.47], p = 0.028; **Figure 4**). Specifically, higher pHPC Glu/NAA was linked to more severe reexperiencing symptoms through stronger pHPC-visual cortex connectivity. In contrast, an alternative model treating pHPC Glu/NAA as the mediator was not significant (indirect effect: B = 0.12, 95% CI = [-0.13, 0.65], p = 0.352).

**Figure 4.**
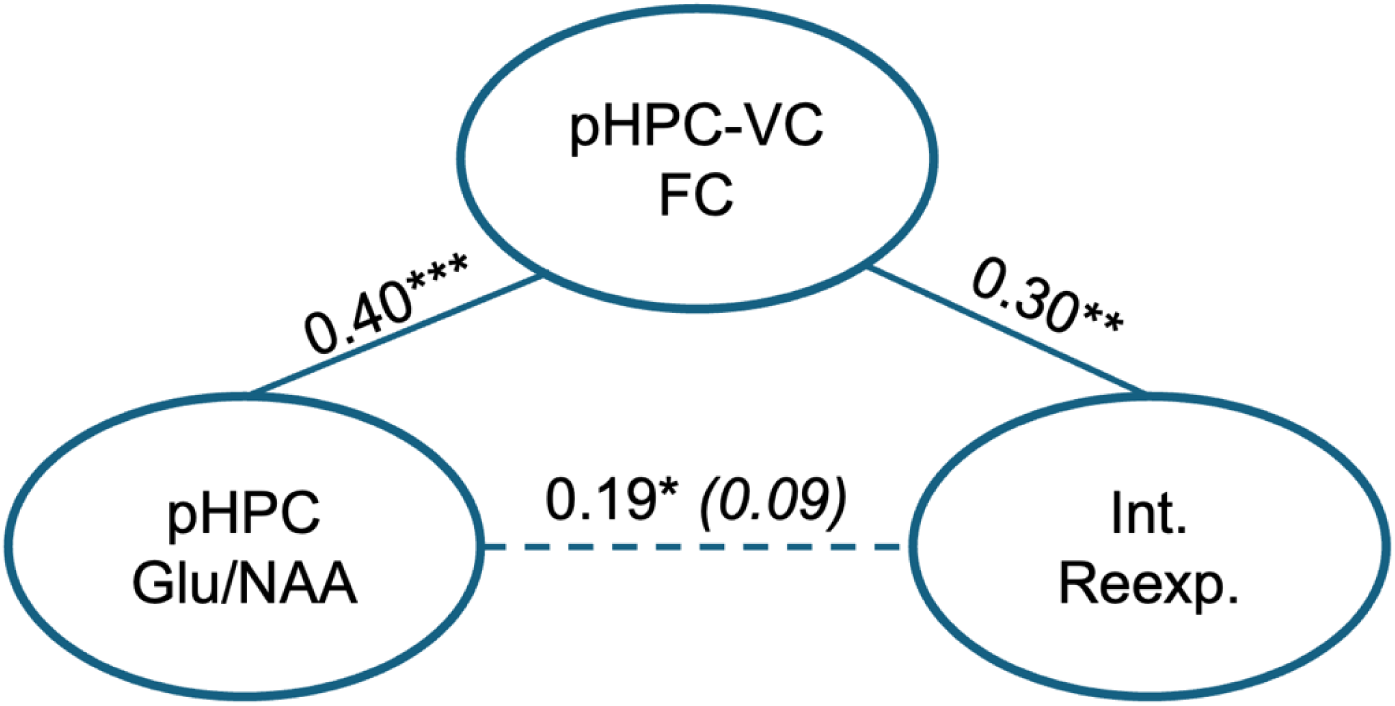
Posterior hippocampus (pHPC)-visual cortex connectivity explained the shared variance between pHPC Glu/NAA and reexperiencing symptom severity. The direct effect between pHPC Glu/NAA and reexperiencing severity (Int. Reexp.) was no longer significant (dashed line) after including functional connectivity between the pHPC and visual cortex (VC) as a mediator. Path strengths are indicated by standardized beta coefficients. The parenthetical beta coefficient reflects the non-significant direct effect after controlling for pHPC-VC FC. \**p* < 0.05; \*\**p* < 0.01; \*\*\**p* < 0.005. Note: Glu = glutamate; NAA = N-acetyl-aspartate.

### Effects of Trauma Load and PTSD Diagnosis

Trauma load was associated with more severe reexperiencing symptoms (sr(105) = 0.25, p = 0.008, 95% CI = [0.07, 0.42]), stronger pHPC-visual cortex connectivity (sr(105) = 0.21, p = 0.031, 95% CI = [0.02, 0.38]), and marginally higher pHPC Glu/NAA (sr(105) = 0.17, p = 0.086, 95% CI = [-0.02, 0.35]). Trauma load did not moderate associations between pHPC Glu/NAA and reexperiencing severity (p = 0.538) or pHPC-visual cortex connectivity (p = 0.685). A marginal moderating effect of trauma load was observed on the association between pHPC-visual cortex connectivity and reexperiencing severity (β = 0.18, SE = 0.10, p = 0.087), with stronger associations at higher trauma loads (**Figure S3**).

There were no significant differences in pHPC Glu/NAA between participants with a PTSD diagnosis and those with subthreshold presentations (t = 1.20, p = 0.237). As expected, the PTSD group reported more severe reexperiencing symptoms (t = 3.33, p = 0.002), and demonstrated a trend towards stronger pHPC-visual cortex connectivity (t = 1.76, p = 0.086). PTSD diagnosis did not moderate associations between reexperiencing symptoms, pHPC Glu/NAA, or pHPC-visual cortex connectivity (p’s > 0.332).

## DISCUSSION

In an independent trauma-exposed sample, this investigation replicates our seminal findings linking higher hippocampal Glu/NAA ratios to more severe intrusive reexperiencing symptoms in PTSD [13]. Additionally, it provides novel evidence that this effect may be specific to the posterior hippocampus. Further, our results identify an association between elevated pHPC Glu/NAA and strengthened pHPC-visual cortex connectivity, which also correlated with reexperiencing severity. Interestingly, stronger connectivity within this pHPC-visual cortex circuit explained the shared variance between pHPC Glu/NAA and reexperiencing severity, offering potential mechanistic insight into the link between hippocampal neurochemistry and memory-based symptoms.

These findings align with excitotoxicity models of hippocampal dysfunction in PTSD, which may be mediated by glutamate [11,32]. Lower hippocampal NAA values, reflecting impaired neuronal integrity, are consistently observed in PTSD, occurring with and without structural atrophy, and covarying with reexperiencing symptoms [13,15]. Although less frequently examined, Glu dysfunction is also implicated in PTSD; however, the direction of dysfunction varies based on the brain region examined. *In vivo* MRS evidence in PTSD demonstrates lower Glu in prefrontal regions that are associated with cognition and emotion, and higher Glu in the hippocampus, temporal cortex, and posterior parieto-occipital regions [15] – structures comprising an episodic memory network [33]. While Glu dysfunction, regardless of directionality or region, is associated with hyperarousal symptoms [15], its link to memory-related reexperiencing symptoms seems to be specific to elevated levels in the hippocampus [13]. Here, our findings corroborate a relationship between hippocampal glutamate metabolism and reexperiencing symptom severity, further suggesting the potential regional specificity of elevated glutamatergic signaling in PTSD to memory-related neural circuits. However, individual Glu and NAA concentrations were not associated with symptom severity in our sample, demonstrating the importance of examining glutamatergic function in the context of other markers of neuronal integrity, as discussed below.

Advancing the idea of regional specificity, we demonstrated here that the association between hippocampal Glu/NAA and intrusive reexperiencing may be specific to the pHPC (versus aHPC). The pHPC is preferentially connected anatomically and functionally to posterior parieto-occipital regions implicated in episodic memory retrieval and detailed mental imagery [18]. By interacting with sensory areas, the pHPC supports vivid sensory details of episodic memories [34], which are core features of reexperiencing symptoms. Conversely, the aHPC, preferentially connected to limbic structures and prefrontal regions associated with emotion regulation and cognition, relates broadly to overall posttraumatic symptom severity [35], and hyperarousal and mood symptoms, but not reexperiencing [24]. Taken together, our findings align with existing evidence implicating pHPC-posterior cortical circuitry in trauma reexperiencing.

We found that greater pHPC Glu/NAA was also linked to stronger functional connectivity between the pHPC and the visual cortex. As the majority of our participants met diagnostic criteria for PTSD, this aligns with evidence of elevated Glu in the hippocampus and parieto-occipital regions in PTSD [13,15], although Glu dysfunction within the visual cortex has been largely unexamined [15]. This lends further credence to the role of glutamatergic dysfunction in a pHPC-posterior cortical circuit involved in memory processes in trauma-related disorders. Greater glutamatergic signaling drives synaptic plasticity and can strengthen coupling between connected or co-active regions [10]. Evidence suggests that local Glu concentrations in network hub regions correlate with stronger functional connectivity within, but not outside, the network [36]. However, under extreme conditions, such as marked excitotoxicity with structural atrophy, elevated Glu or Glu/NAA may disrupt connectivity through reduced synaptic density or neuronal death [32]. Notably, in our sample, Glu/NAA was not associated with pHPC volume (Supplemental Results) – a lack of association consistent with prior findings in severe psychiatric disorders [37,38]. In addition to serving as a proxy for neuronal structural integrity, NAA is also a marker for neuronal metabolism or mitochondrial dysfunction, potentially reflecting bioenergetic functions independent of structural changes [39]. Indeed, prior research has suggested that lower NAA in psychiatric disorders may indicate functional deterioration of neuronal networks without structural neurodegeneration [40]. Therefore, the observed link between elevated Glu/NAA and strengthened connectivity may reflect a state of dysfunctional excitatory neurotransmission that has not reached an atrophic threshold [32]. In the absence of synaptic atrophy, this heightened excitatory state could strengthen connectivity with intrinsically connected regions through glutamatergic neuroplasticity, particularly true if those regions are also disinhibited – as the visual cortex is in PTSD [41].

Alternatively, strengthened connectivity between the pHPC and visual cortex may emerge due to repeated and prolonged activation of trauma memories, a defining characteristic of our sample. Similarly, this repeated hyper-excitation of this memory network could explain elevated Glu/NAA in the pHPC. Future experimental or interventional studies targeting connectivity within this circuit or Glu/NAA within the pHPC are needed to ascertain the mechanisms of these interrelations. Furthermore, evaluating the longitudinal dynamics of pHPC Glu/NAA dysfunction and connectivity in trauma-exposed samples would help determine whether these observed pathophysiologies are risk factors for trauma-related disorders, consequences of trauma exposure, or consequences of repeated intrusive trauma memories.

The observed association between reexperiencing symptoms and strengthened connectivity between the pHPC and visual cortex builds on evidence for an active role of the sensory cortex in threat memories broadly [42,43] and trauma-related memories specifically [44,45]. Models of trauma intrusions propose an exaggerated “sensory representation” of traumatic events, with hyperactivity in primary and associative sensory regions as key neural substrates [23]. Research into the neurobiology of lived trauma experiences highlights the role of the sensory cortex in the visceral nature of reexperiencing symptoms [46]. This aligns with a “sensory disinhibition” model of posttraumatic stress symptoms, proposed by our group and others, wherein hyper-excitability of sensory cortex disrupts resting-state neural network connectivity [41]. Integrating these sensory models with canonical hippocampal models of memory and PTSD, our group recently demonstrated that persistent coactivation between the visual cortex and pHPC (but not aHPC) correlates with the reexperiencing qualities of intrusive trauma memories. Although our cross-sectional mediation model cannot establish causality or directionally, we propose that strengthened connectivity within this circuit may serve as a pathophysiological mechanism linking pHPC Glu/NAA to reexperiencing symptoms. These novel findings further suggest the potential benefit of inhibitory targeting of the visual cortex and its networks for treating trauma reexperiencing. Moreover, the regional specificity of our results to a pHPC-visual circuit supports the use of scalable, visuospatial-based interventions for reexperiencing symptoms [4].

Consistent with prior translational research [10,11], we found that trauma load, indexed by the total number of potentially traumatic events experienced across the lifespan, was associated with elevated pHPC Glu/NAA, albeit weakly. Greater trauma load also correlated with stronger pHPC-visual cortex coupling, consistent with an influence of repeated trauma exposure on hippocampus phenotypes in humans. Interestingly, trauma load weakly moderated the relationship between pHPC-visual cortex connectivity and reexperiencing symptoms, such that this association was absent in individuals with lower trauma exposure. However, trauma load did not moderate relationships with Glu/NAA. Moreover, we did not find any moderating effects of PTSD diagnosis. This suggests that the observed hippocampal neurochemistry is intrinsically linked to the memory-related process of intrusive reexperiencing, regardless of trauma load or PTSD diagnosis. Other features of trauma chronicity, such as developmental timing (i.e., childhood versus adulthood), duration (i.e., multiple acute versus single prolonged), and type, may influence the extent to which trauma load moderates these associations. Additionally, our sample was preferentially recruited for elevated reexperiencing symptoms, which yielded a highly symptomatic sample. Therefore, future studies examining trauma chronicity at a more granular level and including an asymptomatic trauma-exposed control group are warranted.

This study has several limitations to consider. First, as mentioned, the cross-sectional data precludes conclusions about causal relationships among pHPC neurochemistry, connectivity, and reexperiencing symptoms. Second, single voxel MRS methods offer limited spatial resolution - the larger voxel size needed for adequate signal invariably captured portions of brain structures beyond the targeted a/pHPC, potentially introducing partial volume effects. Advances in whole-brain MRS imaging could improve spatial specificity, providing additional evidence for the proposed glutamatergic dysregulation of a posterior memory circuit in reexperiencing symptoms. Relatedly, because our MRS measurements reflect both intracellular and extracellular metabolite concentrations, we cannot distinguish between vesicular and neurotransmitter pools. Finally, more detailed investigations into neuronal integrity and microstructure would help ascertain the extent to which elevated Glu/NAA reflects an excitotoxic mechanism with structural insult or merely heightened excitatory transmission.

In sum, the present findings replicate and extend prior work linking hippocampal excitotoxicity to trauma reexperiencing symptoms by demonstrating a specific effect in the pHPC and its connections to the visual cortex. This identified pHPC-visual cortex circuit integrates clinical models of trauma memory features, historical models of hippocampal dysfunction, and emerging neurobiological models of sensory cortical disinhibition in trauma-related symptoms, specifically intrusive reexperiencing. By gaining a deeper understanding of this pHPC-visual system circuit and its potential molecular underpinnings, we inch closer to developing mechanism-based interventions for the intrusive reexperiencing of trauma. This multi-level, multi-modal approach – spanning molecules, circuits, and symptom phenomenology – may offer useful tools for more personalized and precision-based treatments for trauma survivors.

## Data Availability

All data presented are available through the National Institute of Mental Health Data Archive, and are further available upon reasonable request to the senior author (IMR).

## Acknowledgments

This work was supported by NIH award R01-MH120400 (PI: IMR). Additionally, IMR was partially supported by NIH award P50-MH115874 (Project 4 PIs – IMR, Scott L. Rauch; Program Directors – William A. Carlezon Jr. and Kerry J. Ressler). KJC was supported by NIH award K23-MH137459. The authors would like to thank all participants for dedicating their time and energy to completing the study. We also thank the MRI technologists of the McLean Imaging Center.

