## Supplemental Material for "Multimodal associations between posterior hippocampus glutamate metabolism, visual cortex connectivity, and intrusive trauma reexperiencing symptoms"

### Supplemental Methods

#### *Participants*

Participants were recruited via community advertisements. Inclusion criteria consisted of exposure to a DSM-5 Criterion A trauma and currently experiencing two trauma-related intrusive memories per week within the past month. Exclusion criteria included: left-handedness, medical conditions that could confound results (e.g., uncontrolled neurological disorder), a history of moderate to severe traumatic brain injury, current (past month) moderate-to-severe alcohol or substance use disorder, and current psychotic disorder or manic episode.

#### *Magnetic Resonance Imaging*

Eyes-open resting-state fMRI data were acquired using the HCP Lifespan protocol (T2-weighted echoplanar images; TR/TE: 800/37 ms, in-plane resolution: 2mm; voxels: 2mm isotropic; multiband factor = 8; anterior-posterior phase encoding; one run of 976 frames, ~13 minutes in length). A T1-weighted 3D MPRAGE anatomical image was acquired using the HCP 0.8mm resolution sequence (TR/TEs: 2500/1.81/3.6/5.39/7.18; flip angle: 8 deg; FOV: 256 x 240; voxel size: 0.8mm isotropic). MRI data were preprocessed using fMRIPrep version 20.2.7 [1], followed by additional denoising of white matter and cerebrospinal fluid signals [2], scrubbing of motion outliers (framewise displacement > 0.5 mm) [3]; and high pass (0.01 Hz) filtering using CONN toolbox scripts [4]. Participants were excluded if their mean framewise displacement (FD) exceeded 0.5 mm or greater than 20% of volumes exceeded FD = 0.5mm (n = 10) [5].

Subject-level a/pHPC structural masks were generated using Freesurfer version 7.2.0 [6]. The aHPC was defined as an aggregate of hippocampal subfields within the head region and the pHPC was defined as an aggregate of body and tail regions [7]. Volumes for these aggregates were estimated via Freesurfer and normalized to total intracranial volume. Subject-specific segmentations were then registered to the MNI152 space and merged across participants to create study-specific probabilistic a/pHPC templates for rs-fMRI analyses. We used a template probability threshold of 0.75 to avoid the inclusion of inconsistent voxels. An anterior threshold of y = -32 was used for the pHPC template based on prior work [8]. The resulting a/pHPC masks are depicted in the Supplement Figure S1. Whole-brain seed-to-voxel correlations were performed with the resulting a/pHPC masks to compute whole-brain a/pHPC FC maps using CONN toolbox scripts. Seed-based FC maps were Fisher's z-transformed prior to statistical analyses.

*Magnetic Resonance Spectroscopy*

Single-voxel proton MRS was acquired over 20 x 10 x 20 $mm^3$ voxels positioned in the right posterior and anterior hippocampus. Voxel position and size was optimized to minimize the overlap with temporal cortex white matter and the midline air and CSF spaces. A semi-LASER sequence with VAPOR saturation was used to optimally measure Glu concentrations (TE = 28 ms, TR = 3000 ms, spectral bandwidth = 3.0 kHz, total number of signal averages = 64). B0 shimming was performed using the Siemens FASTESTMAP [9], followed by manual enhancement by a spectroscopist (author XC). Spectra were processed using LCModel with home-simulated basis spectra of metabolites. Spectra were visually inspected for fitting errors by spectroscopists (authors SZ, XS, TS, FD); in addition, a full-width half maximum (FWHM) ≤ 0.05 ppm or (FWHM) ≤ 6 Hz in LCModel and signal-to-noise ratio (SNR) > 15 were required for inclusion in the data analyses. Using these criteria, aHPC data from 7 participants were excluded; no pHPC data were excluded. Metabolite concentrations were referenced to water, controlling for proportions of gray matter, white matter, and cerebrospinal fluid proportions within the MRS voxel and relaxation effects [8,10,11]. Representative spectra are presented in the Supplement (**Figure S2**).

*Statistical Analyses.*

Mediation analyses were performed in R using the *mediate* function [12].

**Supplemental Results**

*Lack of associations with hippocampal volume*

pHPC volume was not associated with pHPC Glu/NAA (r = -0.08, p = 0.415). Similarly, pHPC volume was not associated with reexperiencing symptoms (r = -0.08, p = 0.406) or pHPC-visual cortex connectivity (r = -0.04, p = 0.654). Similarly, there was no association between aHPC volume and aHPC Glu/NAA (r = -0.06, p = 0.525).

*Lack of associations with individual Glu and NAA concentrations*

Reexperiencing symptom severity was not associated with pHPC Glu ($r = 0.17$, $p = 0.069$) or NAA concentrations ($r = -0.013$, $p = 0.885$). Similarly, there was no association with aHPC Glu ($r = 0.17$, $p = 0.073$) or NAA concentrations ($r = 0.16$, $p = 0.100$). Stronger pHPC-visual cortex connectivity was associated with greater pHPC Glu concentrations ($r = 0.23$, $p = 0.018$); however, this effect did not emerge at the whole-brain level, as no clusters survived multiple comparisons correction.

**Supplemental Figures**

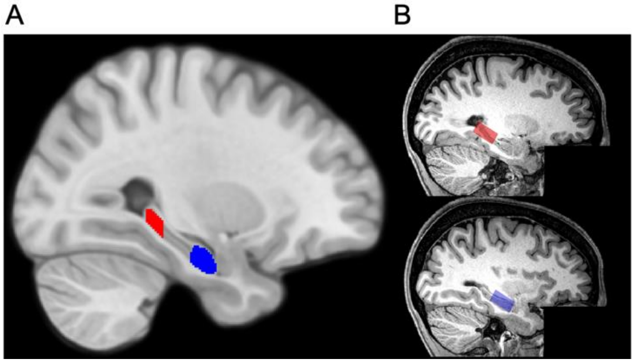


**Figure S1.** Anterior and Posterior Hippocampus Regions of Interest. A) Posterior (red) and anterior (blue) hippocampus masks generated from the study-specific probabilistic template, with a threshold of 0.75, that were used for rs-fMRI analyses. B) Placement of MRS voxels within the posterior (red) and anterior (blue) hippocampus, demonstrating visual overlap with the rs-fMRI masks.

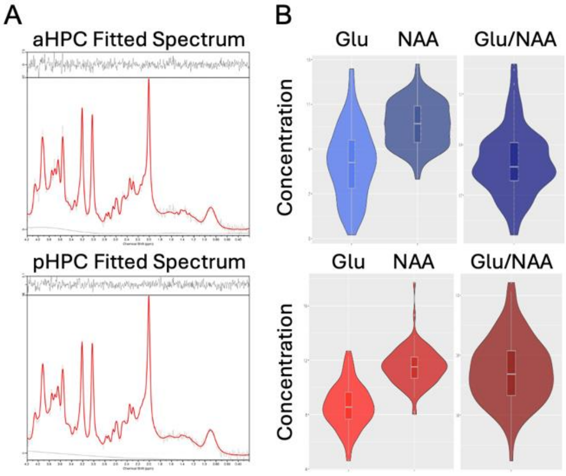


**Figure S2.** Magnetic Resonance Spectroscopy spectra and metabolite concentrations. A) Representative spectra from the aHPC and pHPC voxels. The black spectra represent the raw data, and red spectra represent the model-fitted data, with residuals plotted above. B) Distributions of Glu and NAA concentrations, as well as Glu/NAA ratios.

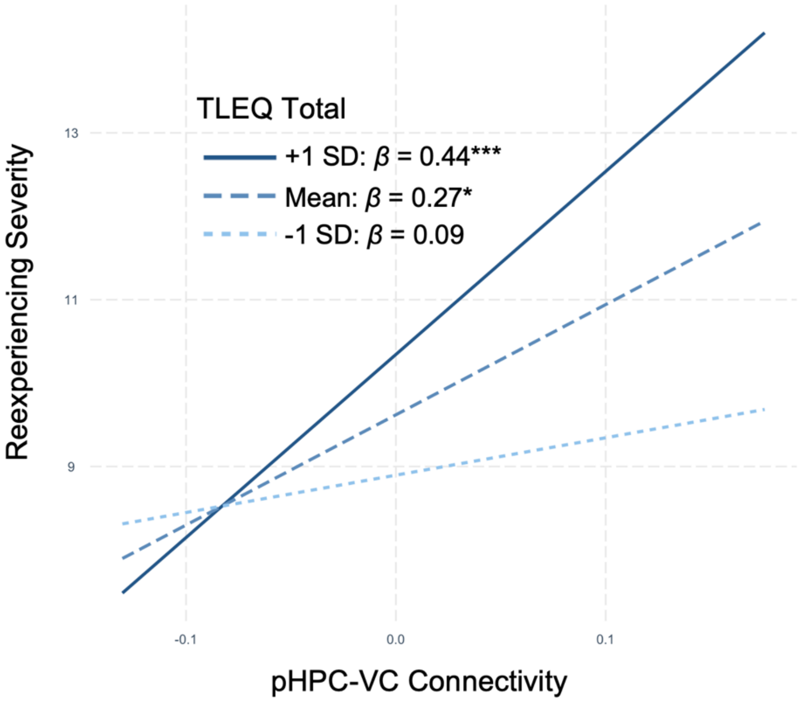

**Figure S3.** TLEQ Trauma Load moderates the association between reexperiencing symptom severity and posterior hippocampus (pHPC)-visual cortex (VC) functional connectivity. Participants who had more total lifetime trauma exposure (+1 SD) demonstrate a positive association (corresponding to a medium effect size) between pHPC-VC connectivity and reexperiencing symptoms, while participants who had less trauma exposure (-1 SD) show no association. However, the interaction of trauma load score with pHPC-VC connectivity was not statistically significant ($p = 0.087$). TLEQ = Traumatic Life Events Questionnaire
